# EPIDEMIOLOGICAL CHARACTERISTICS AND SEVERITY OF OMICRON VARIANT CASES IN THE APHP CRITICAL CARE UNITS

**DOI:** 10.1101/2022.01.25.22269839

**Authors:** Antoine Vieillard-Baron, Rémi Flicoteaux, Maud Salmona, Djillali Annane, Soufia Ayed, Elie Azoulay, Raphael Bellaiche, Sadek Beloucif, Enora Berti, Astrid Bertier, Sébastien Besset, Marlène Bret, Alain Cariou, Christophe Carpentier, Oussama Chaouch, Appoline Chariot, Cyril Charron, Julien Charpentier, Cherifa Cheurfa, Bernard Cholley, Sébastien Clerc, Alain Combes, Benjamin Chousterman, Yves Cohen, Jean-Michel Constantin, Charles Damoisel, Michael Darmon, Vincent Degos, Bertrand De Maupeou D’Ableiges, Sophie Demeret, Etienne De Montmollin, Alexandre Demoule, Francois Depret, Jean-Luc Diehl, Michel Djibré, Chung-Hi Do, Emmanuel Dudoignon, Jacques Duranteau, Muriel Fartoukh, Fabienne Fieux, Etienne Gayat, Mael Gennequin, Bertrand Guidet, Christophe Gutton, Sophie Hamada, Nicholas Heming, Romain Jouffroy, Hawa Keita-Meyer, Olivier Langeron, Brice Lortat-Jacob, Jonathan Marey, Alexandre Mebazaa, Bruno Megarbane, Armand Mekontso-Dessap, Jean-Paul Mira, Julie Molle, Nicolas Mongardon, Philippe Montravers, Capucine Morelot-Panzini, Safaa Nemlaghi, Bao-long Nguyen, Antoine Parrot, Romain Pasqualotto, Nicolas Peron, Lucile Picard, Marc Pineton de Chambrun, Benjamin Planquette, Benoit Plaud, Stéphanie Pons, Christophe Quesnel, Jean-Herlé Raphalen, Keyvan Razazi, Jean-Damien Ricard, Anne Roche, Benjamin Rohaut, Damien Roux, Laurent Savale, Jennifer Sobotka, Jean-Louis Teboul, Jean-François Timsit, Guillaume Voiriot, Emmanuel Weiss, Lucille Wildenberg, Elie Zogheib, Bruno Riou, Frédéric Batteux

## Abstract

**Importance:** Information about the severity of Omicron is scarce.

**Objective:** To report the respective risk of ICU admission in patients hospitalized with Delta and Omicron variants and to compare the characteristics and disease severity of critically ill patients infected with both variants according to vaccination status.

**Design:** Analysis from the APHP database, called Reality, prospectively recording the following information in consecutive patients admitted in the ICU for COVID-19: age, sex, immunosuppression, vaccination, pneumonia, need for invasive mechanical ventilation, time between symptom onset and ICU admission, and in-ICU mortality. Retrospective analysis on an administrative database, “Système d’Information pour le Suivi des Victimes” (SI-VIC), which lists hospitalized COVID-19 patients.

**Setting:** 39 hospitals in the Paris area from APHP group.

**Participants:** Patients hospitalized from December 1, 2021 to January 18, 2022 for COVID-19.

**Main outcomes and measures:** Risk of ICU admission was evaluated in 3761 patients and Omicron cases were compared to Delta cases in the ICU in 888 consecutive patients.

**Results:** On January 18, 45% of patients in the ICU and 63.8% of patients in conventional hospital units were infected with the Omicron variant (p < 0.001). The risk of ICU admission with Omicron was reduced by 64% than with Delta (9.3% versus 25.8% of cases, respectively, p < 0.001). In critically ill patients, 400 had the Delta variant, 229 the Omicron variant, 98 had an uninformative variant screening test and 161 did not have information on variant screening test. 747 patients (84.1%) were admitted for pneumonia. Compared to patients infected with Delta, Omicron patients were more vaccinated (p<0.001), even with 3 doses, more immunocompromised (p<0.001), less admitted for pneumonia (p<0.001), especially when vaccinated (62.1% in vaccinated versus 80.7% in unvaccinated, p<0.001), and less invasively ventilated (p=0.02). Similar results were found in the subgroup of pneumonia but Omicron cases were older. Unadjusted in-ICU mortality did not differ between Omicron and Delta cases, neither in the overall population (20.0% versus 27.9%, p = 0.08), nor in patients with pneumonia (31.6% versus 29.7%, respectively) where adjusted in-ICU mortality did not differ according to the variant (HR 1.43 95%CI [0.89;2.29], p=0.14).

**Conclusion and relevance:** Compared to the Delta variant, the Omicron variant is less likely to result in ICU admission and less likely to be associated with pneumonia. However, when patients with the Omicron variant are admitted for pneumonia, the severity seems similar to that of patients with the Delta variant, with more immunocompromised and vaccinated patients and no difference in adjusted in-ICU mortality. Further studies are needed to confirm our results.

## Introduction

Severe acute respiratory syndrome coronavirus 2 (SARS-CoV-2) infection and the resulting coronavirus disease 2019 (COVID-19) have afflicted millions of people in a worldwide pandemic. A new SARS-CoV-2 variant, B.1.1.529, has recently been reported, with the first cases in Botswana and South Africa on November 11 and 14, 2021, respectively.^1^ On November 26, 2021, B.1.1.529, named Omicron, was designated as a variant of concern by the World Health Organization (WHO).^2^ The Omicron variant shares several mutations with previous Alpha, Beta, and Gamma variants of concern, which immediately raised global concerns about viral transmissibility, pathogenicity, and immune evasion. In the Paris region, the Omicron variant represented 4·1% of daily cases on December 7, but 96·9% of cases 6 weeks later.^3^

Preliminary studies suggested that Omicron is less severe than other variants. Ferguson et al. reported in the UK a 40% reduction in the risk of hospitalization^4^ for Omicron cases, while Wolter et al. recently showed an 80% reduction in South Africa^5^ with no difference in developing severe disease in Omicron cases. In a preliminary report from the electronic administration system of 49 acute care hospitals in South Africa, Maslo et al. suggested that Omicron cases were also less likely to be admitted to the ICU and to receive mechanical ventilation^6^. However, most of these results are based on administrative databases and are strongly related to immune status, including vaccination. Information is still lacking on the characteristics and disease severity of patients admitted to the ICU with the Omicron variant, and on the potential impact of vaccination on disease severity.

All patients admitted to ICUs of the APHP group have been prospectively monitored for COVID-19 since July 2020, leading to a database of more than 7,000 patients. The aim of this study was to compare the characteristics and disease severity of critically ill patients infected with the Omicron and Delta variants according to vaccination status. As APHP also monitors admissions to general medical wards, a further aim was to report the respective risk of ICU admission of patients hospitalized with the Delta and Omicron variants.

## Methods

This study was approved by the ethics and scientific committees of the APHP group (CSE-21-32).

The number of patients admitted to ICUs and to general medical wards was collected on a daily basis from the Système d’Information pour le Suivi des Victimes (SI-VIC) database, which provides real-time data on COVID-19 patients hospitalized in French public and private hospitals (https://www.data.gouv.fr) and was activated for the COVID-19 pandemic on March 13, 2020.^7,8^ As SI-VIC does not provide extensive clinical data on registered patients, in July 2020 we started collecting clinical, vaccinal and virological information from patients admitted to ICUs of the APHP group in the APHP Reality registry.

RT-PCR positivity for SARS-CoV-2 and RT-PCR variant-specific screening tests from patients admitted to ICUs and to general medical wards were collected from the laboratory information system GLIMS® (CliniSys, UK), a software and database shared by all but one virology laboratory of the APHP hospitals. The screening test especially analyzed the following mutations in the SARS-CoV-2 spike protein: L452R (mutation C) and Δ69-70 or N501Y, or K417N (mutation D). Therefore, C1D0 corresponds to a possible Delta variant and C0D1 corresponds to a possible Omicron variant.^9^ Due to the virological distribution of SARS-CoV-2 strains at the time of data analysis, virological results were expressed as Omicron, Delta, or uninformative when screening was impossible, or gave no interpretable results (insufficient viral load), and finally missing. No other variant was observed during the study period.

The Reality database was created in July 2020. It allows intensivists to enter consecutive patients admitted to all the ICUs of APHP group for COVID-19 during the different waves. The following information is collected: age, sex, type of variant, immunosuppression^10^, COVID-19 pneumonia as the reason for admission, time between symptom onset and ICU admission, number of doses of vaccine and the type of vaccine (BNT162b2, mRNA-1273 or other, mostly chAdOx1), invasive mechanical ventilation requirement and finally in-ICU mortality in discharged patients. Only patients with no dose of vaccine were considered unvaccinated, as the natural immunity of patients with 1 dose was unknown.

### Statistical analysis

The respective risk of being admitted to the ICU between December 1, 2021 and January 18, 2022 for Omicron and Delta cases was calculated from the population admitted to hospital for COVID-19 using the administrative database SI-VIC and the laboratory information system GLIMS. Respective characteristics of critically ill Omicron and Delta patients were determined from patients prospectively entered in the Reality registry during the same period.

Categorical data were reported as number and percentage, and medians [Q2, Q3] were reported for continuous data. Information that was not available was treated as missing and no imputation was performed (data were excluded for group comparisons). Percentages were reported as percent of available data (excluding missing values). Comparisons were performed using the Wilcoxon test for continuous variables and the Chi square test for percentages. A survival analysis using a Cox model was performed in the subgroup of critically ill patients with pneumonia including the following variables age, sex, type of variant, immunosuppression, vaccination status and time between symptom onset and ICU admission. The adjusted hazard ratio (aHR) with its 95% confidence interval (95%CI) for the type of variant was reported. A p < 0·05 was considered significant. Statistical analysis was conducted in R (R Core Team 2022).

## Results

### Results from the administrative database: risk of ICU admission

From December 1 to January 18, we identified 5140 patients positive for SARS-CoV-2 by RT-PCR. Among them, 1379 (26.8%) had uninformative variant information and 3761 were positive for either the Delta (1376 patients) or Omicron (2385 patients) variant (**Figure 1**). Among those 3761 patients, 287 (7.6%) were initially admitted to the ICU. Of the 1628 patients initially admitted to a general medical ward, 292 (17.9%) were eventually transferred to the ICU. Among the Delta cases (n=1376), 357 (25.9%) were admitted to the ICU, either initially or after conventional hospitalization. By contrast, in Omicron cases (n=2385) there was a 64% reduction in risk of ICU admission directly or after transfer from the ward (222 patients, 9.3%, p < 0.001 versus Delta). The same significant result was observed among patients hospitalized for more than 1 day on the ward, the risk of transfer to the ICU with Omicron being 51% of that with Delta (20.7% for Omicron *versus* 42.2% for Delta, respectively, p<0.001). Finally, 1315/2385 (55.1%) of Omicron patients finally stayed in the emergency department only and were not admitted to the ICU or to a general medical ward, compared to 531/1376 (38.6%) for Delta patients (p<0.001).

**Figure 1:**
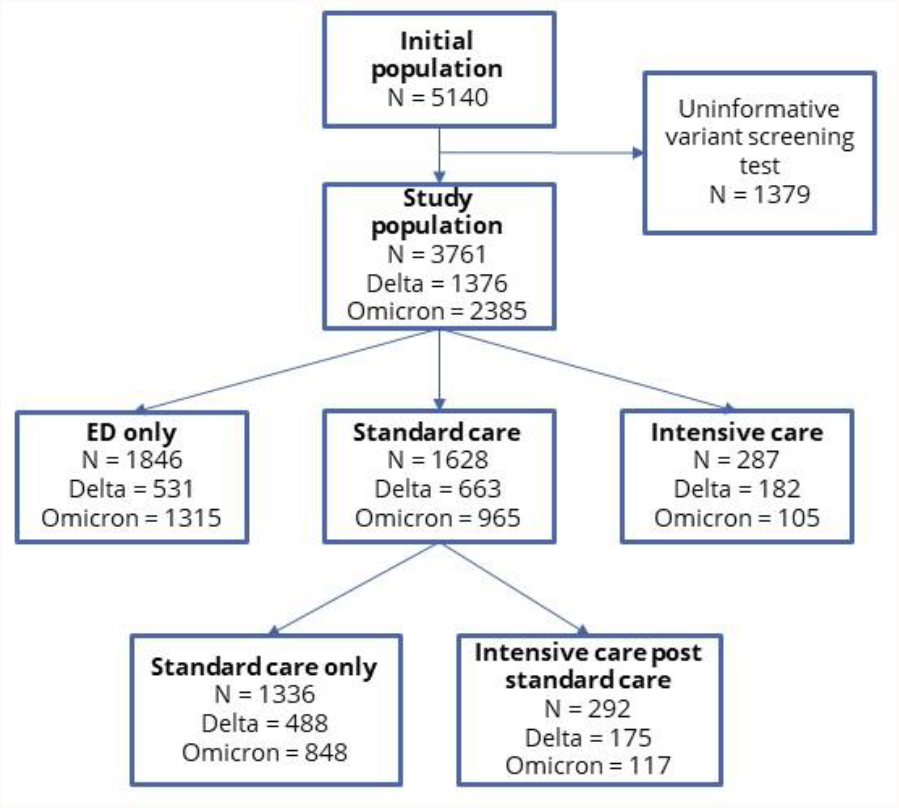
Study flow chart for the 5140 RT-PCR positive patients between December 1, 2021 and January 18, 2022 according to the Omicron and Delta variants and their respective evolution. ED: emergency department.

The respective dynamics of newly admitted patients per day and the overall number of patients admitted to the ICU and to the general medical ward are reported in **Figure 2**. Among patients hospitalized in the ICU for COVID-19 on January 18, 45.4% had an Omicron variant and 37.9% a Delta variant. The respective values for conventional hospitalizations were 63.8% and 10.4% (p < 0.001).

**Figure 2:**
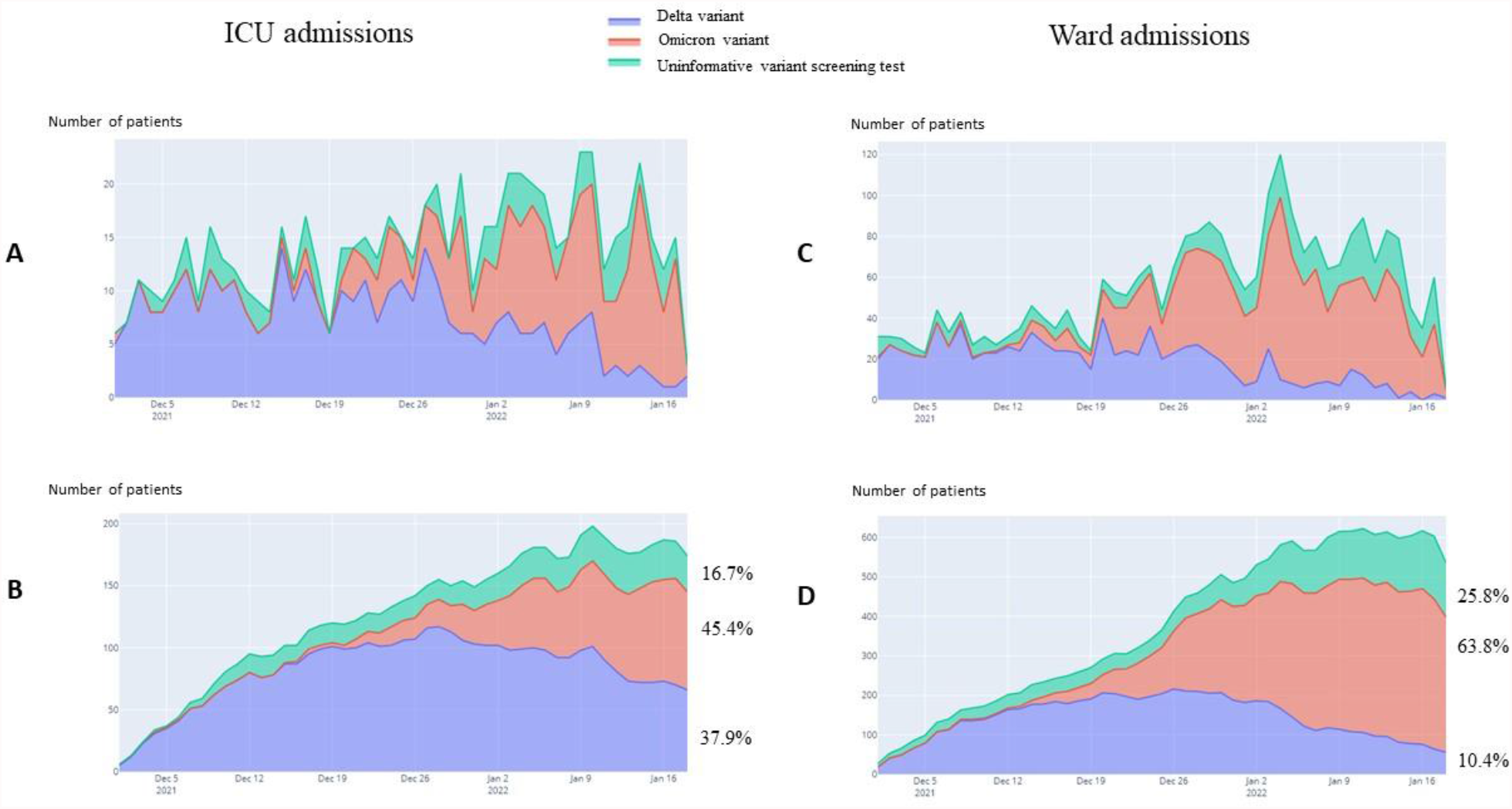
Respective dynamics of the Delta and Omicron variants for new daily ICU admissions (panel A) and the actual number of patients in the ICU during the study period (panel B). Panels C and D report the same information for conventional hospitalization.

### Characteristics and disease severity of critically ill patients with Omicron compared to the Delta variant from the prospective Reality database

As shown in **Table 1**, 888 patients were included in Reality between December 1, 2021 and January 18, 2022. In 161 (18%) patients, we had no variant information. Among the 727 (82%) patients with variant information, 98 (13.5%) had an uninformative variant-screening test (insufficient viral load), 400 (55%) had a Delta variant and 229 (31.5%) had an Omicron variant. Age and sex were similar in Delta and Omicron cases, while with Omicron patients were more immunocompromised (34.5% versus 14.8% for Delta, p < 0.001). No difference was observed for time between symptom onset and ICU admission. Regarding vaccination status, 517 patients (58%) did not receive any injection of vaccine. Omicron infected patients were more frequently vaccinated with at least one injection compared to Delta-positive patients (57.7% versus 30.1%, respectively, p < 0.01). Differences between Delta and Omicron variant were also observed for patients who received 2 or 3 injections (**Table 1**). In patients who had at least 1 injection of vaccine, BNT162b2 or mRNA-1273 was used in 83.5% and other vaccines in 16.5% of cases, mostly chAdOx1 with no difference between the variant.

**Table 1:**
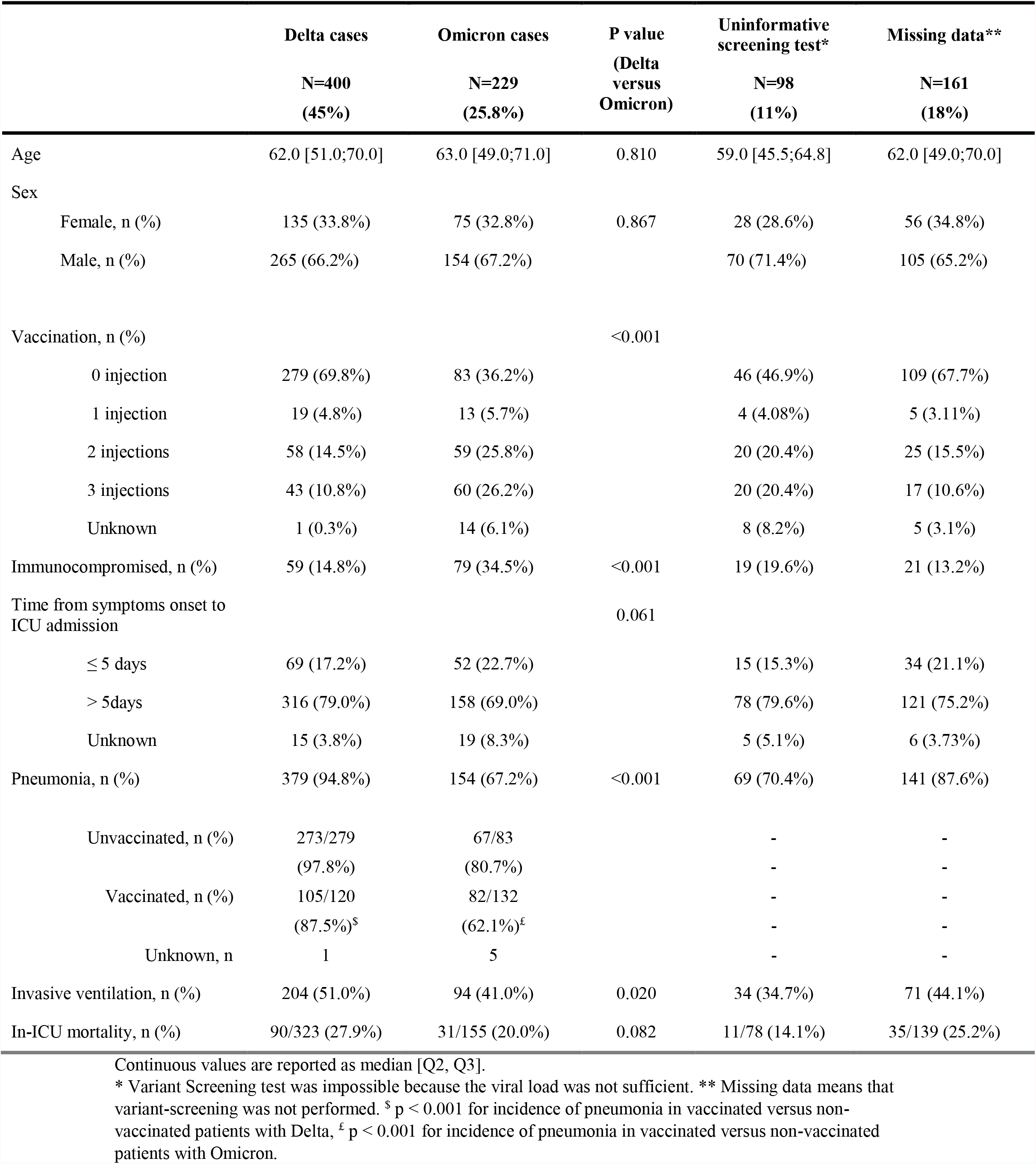
Characteristics of critically ill patients according to Omicron versus Delta variant in the 888 patients included in the Reality database from December 1, 2021 to January 18, 2022.

|  | Delta cases<br>N=400<br>(45%) | Omicron cases<br>N=229<br>(25.8%) | P value<br>(Delta<br>versus<br>Omicron) | Uninformative<br>screening test*<br>N=98<br>(11%) | Missing data**<br>N=161<br>(18%) |
| --- | --- | --- | --- | --- | --- |
| Age | 62.0 [51.0;70.0] | 63.0 [49.0;71.0] | 0.810 | 59.0 [45.5;64.8] | 62.0 [49.0;70.0] |
| Sex |  |  |  |  |  |
| Female, n (%) | 135 (33.8%) | 75 (32.8%) | 0.867 | 28 (28.6%) | 56 (34.8%) |
| Male, n (%) | 265 (66.2%) | 154 (67.2%) |  | 70 (71.4%) | 105 (65.2%) |
| Vaccination, n (%) |  |  | <0.001 |  |  |
| 0 injection | 279 (69.8%) | 83 (36.2%) |  | 46 (46.9%) | 109 (67.7%) |
| 1 injection | 19 (4.8%) | 13 (5.7%) |  | 4 (4.08%) | 5 (3.11%) |
| 2 injections | 58 (14.5%) | 59 (25.8%) |  | 20 (20.4%) | 25 (15.5%) |
| 3 injections | 43 (10.8%) | 60 (26.2%) |  | 20 (20.4%) | 17 (10.6%) |
| Unknown | 1 (0.3%) | 14 (6.1%) |  | 8 (8.2%) | 5 (3.1%) |
| Immunocompromised, n (%) | 59 (14.8%) | 79 (34.5%) | <0.001 | 19 (19.6%) | 21 (13.2%) |
| Time from symptoms onset to<br>ICU admission |  |  | 0.061 |  |  |
| ≤ 5 days | 69 (17.2%) | 52 (22.7%) |  | 15 (15.3%) | 34 (21.1%) |
| > 5days | 316 (79.0%) | 158 (69.0%) |  | 78 (79.6%) | 121 (75.2%) |
| Unknown | 15 (3.8%) | 19 (8.3%) |  | 5 (5.1%) | 6 (3.73%) |
| Pneumonia, n (%) | 379 (94.8%) | 154 (67.2%) | <0.001 | 69 (70.4%) | 141 (87.6%) |
| Unvaccinated, n (%) | 273/279<br>(97.8%) | 67/83<br>(80.7%) |  | -<br>- | -<br>- |
| Vaccinated, n (%) | 105/120<br>(87.5%)<sup>\$</sup> | 82/132<br>(62.1%)<sup>£</sup> | | -<br>- | -<br>- |
| Unknown, n | 1 | 5 |  | - | - |
| Invasive ventilation, n (%) | 204 (51.0%) | 94 (41.0%) | 0.020 | 34 (34.7%) | 71 (44.1%) |
| In-ICU mortality, n (%) | 90/323 (27.9%) | 31/155 (20.0%) | 0.082 | 11/78 (14.1%) | 35/139 (25.2%) |

Continuous values are reported as median [Q2, Q3].
\* Variant Screening test was impossible because the viral load was not sufficient. \*\* Missing data means that variant-screening was not performed. <sup>\$</sup> p < 0.001 for incidence of pneumonia in vaccinated versus non-vaccinated patients with Delta, <sup>£</sup> p < 0.001 for incidence of pneumonia in vaccinated versus non-vaccinated patients with Omicron.

743 patients (83.7%) were admitted to the ICU for pneumonia, while such admission was less frequent for Omicron cases (67.2% versus 94.8% for Delta, p < 0.001, risk reduction of 29%, **Table 1**). In Omicron infected patients, pneumonia was significantly less frequent in vaccinated than in unvaccinated patients (62.1 versus 80.7%, p < 0.001), while among the 60 Omicron infected patients who had 3 doses of vaccine, 43 (71.7%) were admitted for pneumonia. A similar result, even less pronounced, was observed for Delta cases (p < 0.01 for vaccinated versus unvaccinated patients, **Table 1**). Finally, Omicron cases were less frequently invasively ventilated compared to Delta cases (41.0% versus 51%, respectively, p = 0.02). Among patients already discharged from the ICU on January 18, 2021 (80% of Delta cases and 63.6% of Omicron cases), unadjusted in-ICU mortality did not differ (20.0% and 27.9% for Omicron and Delta, respectively, p = 0.08).

In the subgroup of 743 patients admitted to the ICU with pneumonia, 379 had the Delta variant and 154 the Omicron variant (**Table 2)**. Omicron patients were significantly older (see also **Figure 3**), more frequently admitted to the ICU in the 5 days following symptom onset (p = 0.004) and less likely to be invasively ventilated (39% versus 50.4% for Delta, p 0.021). In-ICU mortality did not differ in the 75% of patients already discharged on January 18, 2022 (80% of Delta cases were discharged and 63% of Omicron cases), and aHR for the type of variant was non-significant (1.43, 95%CI [0.89;2.29], p=0.14).

**Table 2:** Characteristics of critically ill patients according to Omicron versus Delta variant in the subgroups of 743 patients admitted in ICU for pneumonia from December 1, 2021 to January 18, 2022.

|  | Delta cases<br>N=379 | Omicron cases<br>N=154 | P value<br>(Delta<br>versus<br>Omicron) | Uninformative<br>screening test*<br>N=69 | Missing data**<br>N=141 |
| --- | --- | --- | --- | --- | --- |
| Age | 62.0 [51.0;70.0] | 65.0 [56.0;72.0] | 0.033 | 60.0 [56.0;65.0] | 62.0 [50.0;70.0] |
| Female, n (%) | 126 (33.2%) | 56 (36.4%) | 0.557 | 20 (29.0%) | 49 (34.8%) |
| Immunocompromised,<br>n (%) | 56 (14.8%) | 61 (39.6%) | <0.001 | 16 (23.2%) | 17 (12.0%) |
| Time from symptoms onset to<br>ICU admission, n (%) |  |  | 0.004 |  |  |
| ≤ 5 days | 61 (16.1%) | 42 (27.3%) |  | 12 (17.4%) | 30 (21.3%) |
| > 5days | 303 (80.0%) | 105 (68.2%) |  | 56 (81.2%) | 107 (75.9%) |
| Unknown | 15 (4.0%) | 7 (4.5%) |  | 1 (1.4%) | 4 (2.8%) |
| Invasive ventilation, n<br>(%) | 191 (50.4%) | 60 (39.0%) | 0.021 | 23 (33.3%) | 62 (44.0%) |
| In-ICU mortality, n (%) | 90/303 (29.7%) | 31/98 (31.6%) | 0.780 | 10/54 (18.5%) | 35/120 (29.2%) |

Continuous values are reported as median [Q2, Q3].
\* Variant Screening test was impossible because the viral load was not sufficient. \*\* Missing data means that variant-screening was not performed.
In-ICU mortality was calculated among the 303 Delta patients (80%) and the 98 Omicron patients (63.6%) who were already discharged from the ICU.

**Figure 3:**
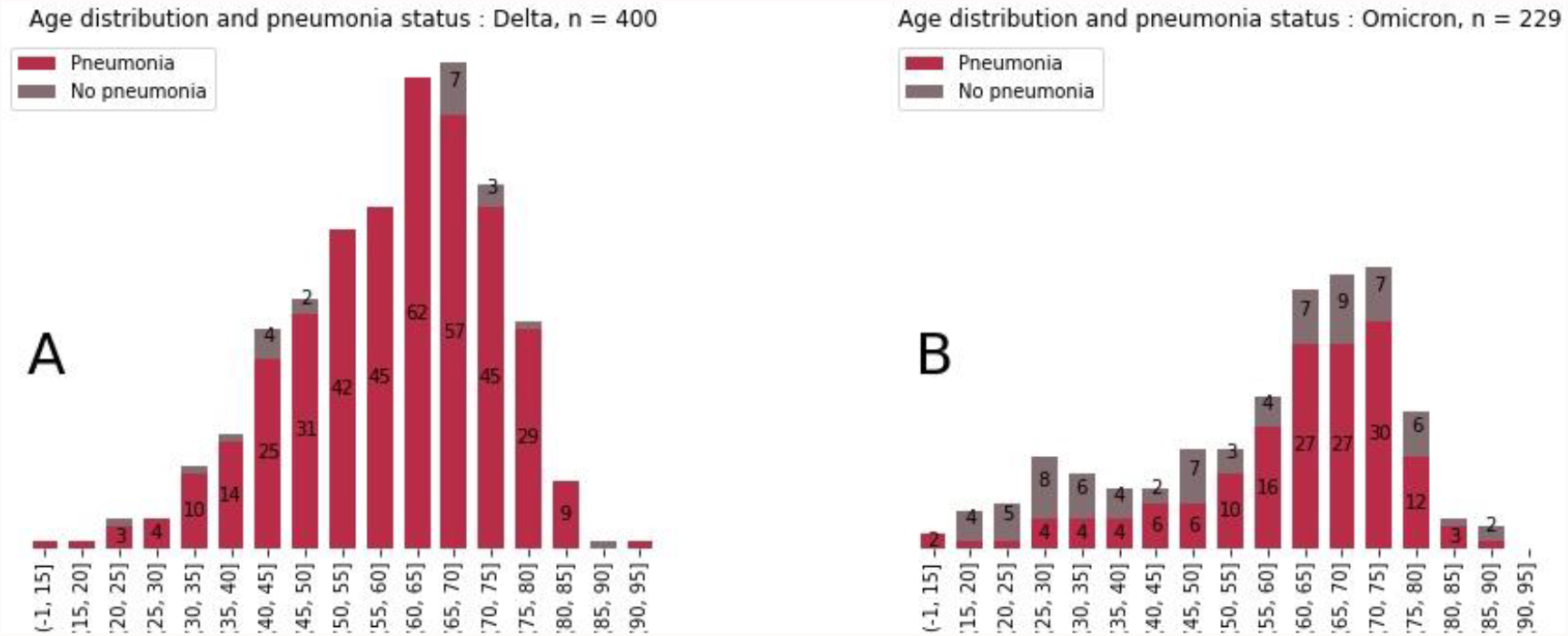
Proportion of pneumonia cases in critically ill patients with the Delta and Omicron variant according to age.

## Discussion

We report the impact of the Omicron variant on ICU admission in patients with COVID-19 hospitalized in the APHP group between December 1, 2021 and January 18, 2022. Omicron patients were 64% less likely to be admitted to the ICU than Delta patients, and more than one half of Omicron infected patients stayed in the emergency department, compared to 39% of Delta infected patients. The rate of increase in patients admitted to the ICU and to the general medical ward showed a faster impact of Omicron on conventional hospitalization than on ICU admission, and Omicron tended to replace Delta more rapidly in the general medical ward than in the ICU. Such results were already suggested from observations in the UK and South Africa.^4,5^ On the other hand, we are able to report that almost one-third of patients hospitalized in the ICU of the APHP group between December 1, 2020 and January 18, 2021 already had the Omicron variant, with a rate of 45% on January 18. It is also noteworthy that patients infected with the Omicron variant were more frequently vaccinated than those with the Delta variant, mostly with an mRNA vaccine for both, even with the booster, which could suggest a lower efficacy of the vaccine. However, they were more immunocompromised and had a 29% reduction in risk of being admitted for COVID-19 pneumonia when they were vaccinated, which could suggest moderate protection of vaccination for the most severe form reported in ICU in unvaccinated patients.^11^ Admissions not due to pneumonia may be related to several causes, as patients admitted to the ICU with a positive RT-PCR but not for COVID-19, severe decompensation of a chronic disease due to direct SARS-CoV-2 infection or another disease pattern related to this new variant. It has been reported that Omicron replicates 10 times less in human lung tissue than the wild strain.^12^ Unfortunately, the design of our registry did not allow us to go further in the analysis.

On January 7, 2022, the Intensive Care National Audit and Research Center (ICNARC) reported epidemiological data about critically ill COVID-19 patients.^13^ There was a dissociation between the number of hospital admissions (significant increase) and in-ICU admission (quite stable) since December 2021. However, the comparison relates to two long periods of time, the first between September 1, 2020 and April 30, 2021, the second between May 1 2021 and January 7 2022, which makes it difficult to draw any conclusion regarding the new Omicron, which that emerged in December 2021. Above all, no information was given about the type of variant and so it was not possible to compare Delta and Omicron cases.^13^

With our prospective Reality database, we are able to provide new information on the comparison between Delta and Omicron cases. Besides the difference in vaccination status discussed above, Omicron patients were more frequently immunocompromised and less frequently invasively ventilated. Unadjusted in-ICU mortality was not different in the global population. More interestingly, among the subgroup of patients admitted to the ICU for pneumonia only, Omicron cases seem to be as severe as Delta cases, with no difference observed in the adjusted risk of mortality. However, Omicron patients were still less invasively ventilated than Delta cases, which could be explained by still more immunocompromised patients.^10^ Among patients admitted for pneumonia, we noted a relatively low rate of invasive ventilation (40-50% of cases), which could be explained first because ICUs in the APHP group usually include a mix of full ICU beds and of intermediate beds for less critically ill patients, and second because compared to the first wave, some studies have now suggested the beneficial effect of high-flow oxygen saturation^14,15^ and non-invasive ventilation^16^ on the requirement for invasive ventilation and on recovery, which probably led to changes in practices.

Our study has some limitations. First, there was a discrepancy between the numbers of critically ill patients with the Delta or Omicron variant in the Reality registry (629 patients) and in the administrative databases (579 patients). As noted in the methods section, the explanation is that all but one hospital of the APHP group collected patient information in the laboratory information system. In Reality, variant screening test information was missing in 18% of cases, which could be explained by the fact that numerous patients were transferred to the ICU from another non-APHP hospital where they had already tested positive. As the test was not performed again we were not able to characterize the variant. Second, we did not have the time between the last injection of vaccine and admission to the ICU, which could limit our evaluation of the impact of vaccination on the Omicron variant. Third, in the adjusted risk of in-ICU mortality evaluation, some confounding factors may not have been taken into account and we do not have information on “do-not-intubate” decision in some patients. Finally, while to our knowledge we report here the largest cohort of critically ill patients comparing Delta and Omicron variant, our results should be confirmed in a larger cohort. A significant number of patients are still in the ICU, especially with the Omicron variant, which precludes any definitive conclusion regarding their respective outcomes.^17^

In conclusion, our study reports contrasting results. On the one hand, Omicron infected patients are less likely to be admitted to the ICU and when admitted, less often for pneumonia. On the other hand, vaccination even with 3 doses had a moderate protective effect against admission for pneumonia in patients who were more immunocompromised. Moreover, when admitted to the ICU for pneumonia, disease severity appears to be similar to that of Delta, with no difference in the adjusted risk of in-ICU mortality. Further studies are needed to evaluate its outcome.

## Data Availability

All data produced in the present work are contained in the manuscript.

## Author contributions

All authors contributed, directly or indirectly, to entering critically ill patients in the Reality database.

A Vieillard-Baron and F Batteux created the Reality prospective registry, analyzed the results and wrote the manuscript.

A Vieillard-Baron designed the study.

F Batteux, R Flicoteaux and B De Maupeou D’Ableiges performed the statistical analysis.

All authors confirm that they had full access to all of the study data and accept responsibility to submit for publication. They all agree with the manuscript.

## Acknowledgments

We thank the “Direction de la recherche clinique et de l’innovation (DRCI)” of APHP group for providing extensive data.

We thank Guillermo Hayoun from the “Direction des systèmes de l’information” for his work in designing and updating the Reality registry when necessary.

We thank Laure Maillant of the health database of APHP (“Entrepôt de Données en Santé”) for her help in generating the data.

Finally, we also thank all the virology laboratories of the APHP group, which helped with the RT-PCR screening of our patients.

